# Integration of pharmacists’ knowledge into a predictive model for teicoplanin dose planning

**DOI:** 10.1101/2023.12.14.23299934

**Authors:** Tetsuo Matsuzaki, Tsuyoshi Nakai, Yoshiaki Kato, Kiyofumi Yamada, Tetsuya Yagi

**Author notes:** Corresponding author: Tetsuya Yagi, Department of Infectious Diseases, Nagoya University Hospital, Nagoya, Aichi, 466-8560, Japan.

## Abstract

Teicoplanin is an important antimicrobial agent for methicillin-resistant *Staphylococcus aureus* infections. To enhance its clinical effectiveness while preventing adverse effects, therapeutic drug monitoring (TDM) of teicoplanin trough concentration is recommended. Given the importance of the early achievement of therapeutic concentrations for treatment success, initial dosing regimens are deliberately designed based on patient information.

Considerable effort has been dedicated to developing an optimal initial dose plan for specific populations; however, comprehensive strategies for tailoring teicoplanin dosing have not been successfully implemented. The initial dose planning of teicoplanin is conducted at the clinician’s discretion and is thus strongly dependent on the clinician’s experience and expertise.

The present study aimed to use a machine learning (ML) approach to integrate clinicians’ knowledge into a predictive model for initial teicoplanin dose planning. We first confirmed that dose planning by pharmacists dedicated to TDM (hereafter TDM pharmacists) significantly improved early therapeutic target attainment for patients without an intensive care unit or high care unit stay, providing the first evidence that dose planning of teicoplanin by experienced clinicians enhances early teicoplanin therapeutic exposure. Next, we used a dataset of teicoplanin initial dose planning by TDM pharmacists to train and implement the model, yielding a model that emulated TDM pharmacists’ decision-making for dosing. We further applied ML to cases without TDM pharmacist dose planning and found that the target attainment rate of the initial teicoplanin concentration markedly increased. Our study opens a new avenue for tailoring the initial dosing regimens of teicoplanin using a TDM pharmacist-trained ML system.

**Importance:** Teicoplanin is used for treating methicillin-resistant *Staphylococcus aureus* infections. Given the importance of early adequate teicoplanin exposure, initial dosing regimens are adjusted for patient characteristics. However, tailoring teicoplanin dosing is challenging for most clinicians. In this study, we first showed that initial dosing regimens by pharmacists dedicated to therapeutic drug monitoring significantly improved early achievement of targeted concentration. In addition, we leveraged machine learning approach to develop the predictive model that tailors initial dosing regimens at the levels of experienced pharmacists. The target attainment rate of patients without experienced pharmacists’ dose planning was significantly increased by applying the model. Therefore, machine learning approach may provide new avenues for tailoring initial teicoplanin dosing.

## 1. Introduction

Teicoplanin is a glycopeptide antibiotic with clinical efficacy in the treatment of methicillin-resistant *Staphylococcus aureus* (MRSA) infections, along with vancomycin (1, 2). With a growing body of evidence supporting the relationship between teicoplanin exposure and response, therapeutic drug monitoring (TDM) is routinely used to maximize clinical effectiveness while preventing adverse effects, such as nephrotoxicity and thrombocytopenia (3, 4). Trough concentration of teicoplanin is considered a key predictor of its effectiveness and adverse effects: trough levels of 15–30 mg/L are recommended for the treatment of noncomplicated MRSA infections, whereas trough levels of 20–40 mg/L have been recently suggested for serious and/or complicated MRSA infections, such as endocarditis and osteomyelitis (2).

Initial teicoplanin dosing starts with multiple loading doses (three to four times, in general), followed by a series of maintenance doses (2, 5, 6). Loading and maintenance doses are adjusted for patient characteristics, such as age, sex, body weight (BW), body mass index (BMI), serum albumin level, and renal function. Given the importance of the early achievement of therapeutic exposure for clinical success, numerous efforts have been made to implement initial dosing nomograms to achieve early attainment of therapeutic concentration levels (5–7). However, because these nomograms were developed and validated for a specific population, their robustness against population changes is limited. To date, the dosing nomograms for individually optimized initial dosing remain controversial, which poses a challenge for the initial dose planning of teicoplanin. In clinical settings, decision-making for initial teicoplanin dosing regimens is often based on the clinician’s experience and expertise (hereafter knowledge) (8–10).

Machine learning (ML), a type of artificial intelligence, provides a set of tools that improve the discovery and decision-making for specific questions with abundant and multidimensional data. A growing body of evidence suggests that ML is a promising approach for medical research and clinical care (11, 12). Recently, ML approaches have been adopted to facilitate TDM studies. Imai et al. developed a nomogram for an initial vancomycin dosing regimen by integrating a dataset of patients treated with vancomycin into ML (13, 14). We previously adopted imitation learning, which is an ML technique that leverages expert demonstrations to learn policies (15). In this study, we defined experts as pharmacists who were experienced in TDM practice and used the dataset of the initial dosing regimen designed by experts to integrate experts’ knowledge into the ML model (16). This straightforward approach has yielded a predictive model that designs initial dosing regimens akin to those of pharmacists, exemplifying the potential of ML techniques for integrating expert knowledge into the model.

In the present study, we aimed to extend this approach to teicoplanin. Using a dataset of dose planning by pharmacists dedicated to TDM practice (hereafter TDM pharmacist), we trained ML to predict the TDM pharmacists’ initial dose planning of teicoplanin. This approach achieved a predictive model that was comparable to that of TDM pharmacists for target trough attainment. In addition, the target attainment rate of patients without TDM pharmacists’ dose planning would significantly increase by applying ML. Our study highlights the clinical significance of integrating pharmacist knowledge into predictive models using ML techniques.

## 2. Results

### 2.1 Patients’ characteristics

We enrolled patients who received teicoplanin between August 2019 and April 2022 at Nagoya University Hospital. During the study period, 1165 patients received teicoplanin treatment. Of these, 751 patients were excluded because of the following reasons: age <18 years (n = 546), undergoing peritoneal dialysis or hemodialysis (n = 109), receiving TDM pharmacists’ intervention after the initial dose (n = 33), resuming teicoplanin treatment within 7 days (n = 31), on extracorporeal membrane oxygenation (ECMO) (n = 16), missing data (n = 9), patient immobility (n = 6), or receiving a single dose for surgical prophylaxis (n = 1). The remaining 414 patients were divided into two groups based on whether they received TDM pharmacist intervention (intervention group) or not (nonintervention group). Consequently, 158 and 256 patients were assigned to the intervention and nonintervention groups, respectively (Figure 1 and Tables S1 and S2).

**Figure 1.**
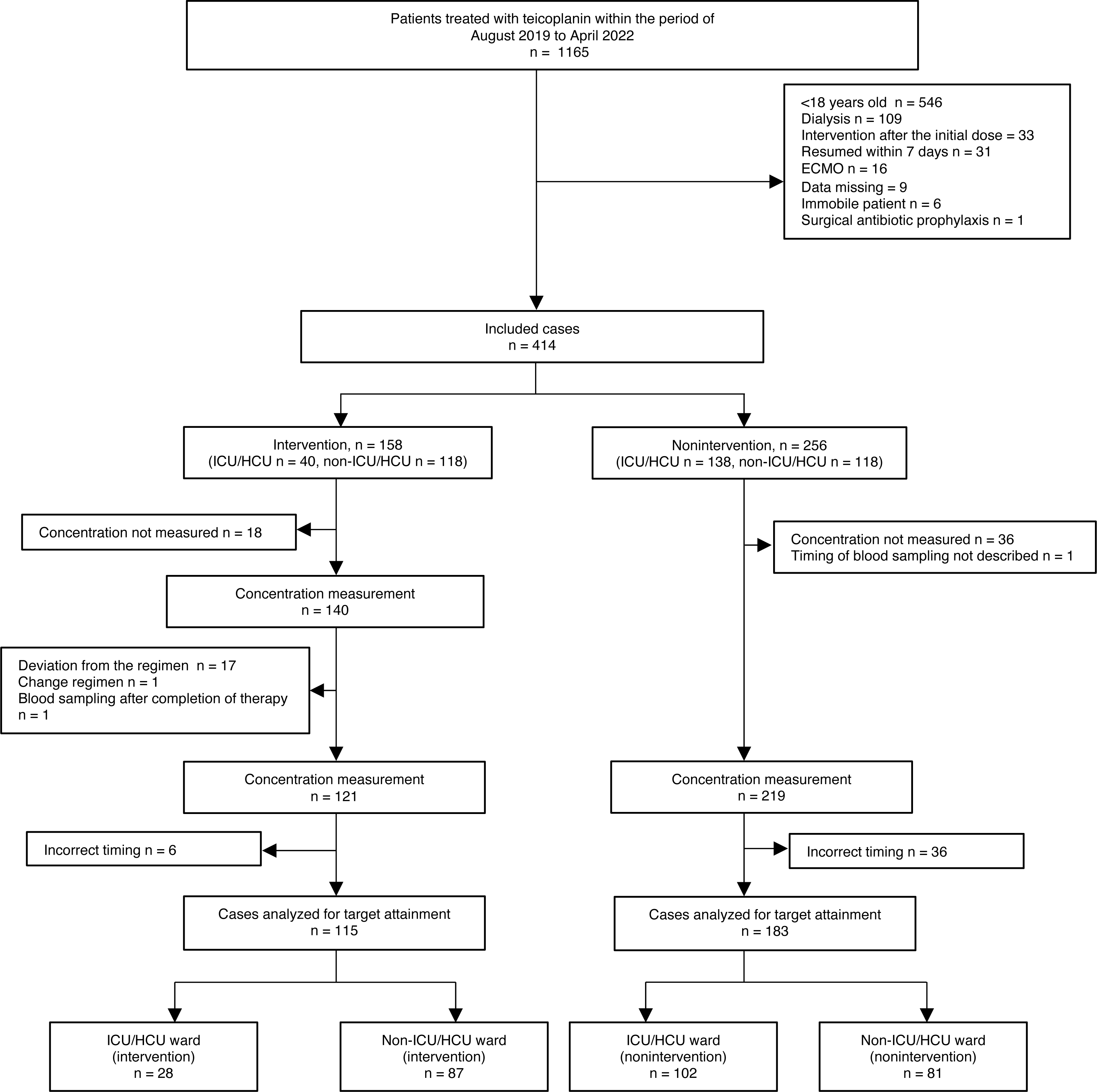
Flow of case selection in the study.

### 2.2 Evaluation of therapeutic drug monitoring pharmacists’ dose planning

We first evaluated the clinical significance of TDM pharmacists’ intervention in teicoplanin treatment. Eighteen of the 158 patients in the intervention group and 37 of the 256 patients in the nonintervention group were excluded from the analysis because of the lack of concentration measurement or description of the sampling time. For the remaining 140 patients in the intervention group, we further excluded the patients with deviation from the TDM pharmacists’ regimen (n = 17), changing regimen after the initial dose (n = 1), and blood sampling after the completion of therapy (n = 1).

For the remaining 121 and 219 patients in the intervention and nonintervention groups, respectively, we assessed whether blood samples were collected at appropriate time points, that is, 18 h after the last dose (2). The incidence rates of inappropriate blood sampling were 5.0% (6/121) and 16.8% (36/219) in the intervention and nonintervention groups, respectively, indicating the contribution of TDM pharmacists’ intervention to proper trough blood sampling (Figure 1 and Table 1). Patients with inappropriate blood sampling in the intervention group (n = 6) were due to the lack of TDM pharmacists’ recommendation on the timing for sampling.

**Table 1.** Blood sampling time.

| Group<br>Sampling time | Intervention (%) | Nonintervention (%) | p-value<br>(Fisher's exact test) |
| --- | --- | --- | --- |
| Correct | 115 (95.0%) | 183 (83.6%) | 0.0017 |
| Incorrect | 6 (5.0%) | 36 (16.4%) |  |

Next, we excluded these patients with inappropriate blood sampling and then analyzed whether TDM pharmacists’ dose planning contributes to target attainment (15–30 mg/L). As shown in Table 2, TDM pharmacist intervention clearly increased the target attainment rate at initial TDM (74.8% [86/115] in the intervention group vs. 57.9% [106/183] in the nonintervention group, p = 0.004), accompanied by a decrease in subtherapeutic ranges (14.8% [17/115] in the intervention group vs. 37.2% [68/183] in the nonintervention group, Table 2 and Tables S3 and S4). However, we observed a large bias in intensive care unit (ICU) and high care unit (HCU) admissions between the two groups (24.3% [28/115] in the intervention group vs. 55.7% [102/183] in the nonintervention group, p <1.0 × 10^−6^, Table S5). Thus, we performed subgroup analyses and found no significant increase in target attainment in patients with an ICU/HCU stay (60.7% [17/28] in the intervention group vs. 57.8% [59/102] in the nonintervention group, p = 0.832, Table 2). This reflects the difficulty in predicting the pharmacokinetics of ICU/HCU stay, where frequent and dramatic changes in the patients’ clinical status are often observed. For patients without an ICU/HCU stay, we found a marked increase in the target attainment rate in the intervention group (79.3% [69/87] in the intervention group vs. 58.0% [47/81] in the nonintervention group, p = 0.004, Table 2). Because there were systemic differences in baseline characteristics between the two groups, especially creatinine clearance (CL_CR_) (p = 0.009, Figure S1 and Table 3), we applied propensity score matching to reduce the effects of confounding. The results showed that the increase was also found and remained significant after propensity score matching (83.3% [40/48] in the intervention group vs. 60.4% [29/48] in the nonintervention group, p = 0.013, Table 4 and Table S6).

**Table 2.** Target attainment.

|  | Intervention |  |  | Nonintervention |  |  | p-value<br>(Chi-square test) |
| --- | --- | --- | --- | --- | --- | --- | --- |
| Trough (mg/L)<br>Group | <15 | 15–30 | >30 | <15 | 15–30 | >30 |  |
| Whole (%) | 17 (14.8) | 86 (74.8) | 12 (10.4) | 68 (37.2) | 106 (57.9) | 9 (4.9) | 0.004 |
| ICU/HCU (%) | 9 (32.1) | 17 (60.7) | 2 (7.1) | 38 (37.3) | 59 (57.8) | 5 (4.9) | 0.832 |
| non-ICU/HCU (%) | 8 (9.2) | 69 (79.3) | 10 (11.5) | 30 (37.0) | 47 (58.0) | 4 (4.9) | 0.004 |

Abbreviations: ICU, intensive care unit; HCU, high care unit.

**Table 3.**
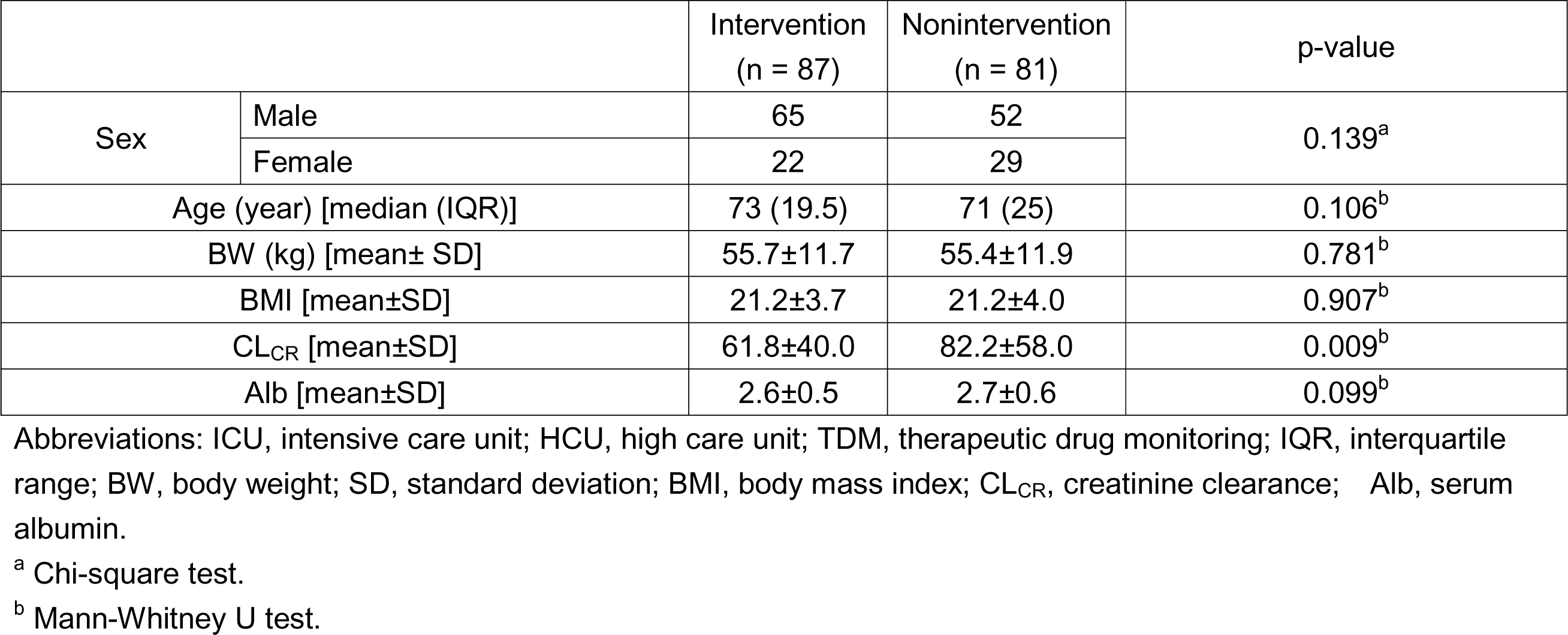
Demographic, clinical, and laboratory characteristics of eligible cases without ICU/HCU admission for TDM analysis.

**Table 4.** Target attainment after propensity score matching (non-ICU/HCU)

| Group<br>Trough (mg/L) | Intervention (%) | Nonintervention (%) | p-value<br>(Chi-square test) |
| --- | --- | --- | --- |
| <15 | 5 (10.4) | 16 (33.3) | 0.013 |
| 15–30 | 40 (83.3) | 29 (60.4) |  |
| >30 | 3 (6.3) | 3 (6.3) |  |

Abbreviations: ICU, intensive care unit; HCU, high care unit.

Overall, these results validate the TDM pharmacists’ dose planning, at least for patients without ICU/HCU stay, and underscore the importance of experts’ (e.g., TDM pharmacists) intervention for effective and appropriate initial treatment with teicoplanin.

### 2.3 Machine learning (ML) model to determine teicoplanin initial dose

Given the increased target achievement by TDM pharmacists’ dose planning of teicoplanin for patients without an ICU/HCU stay, we next sought to build an ML model that tailored dosing regimens at the levels of TDM pharmacists. Toward this end, we trained a two-layer neural network with the dataset of TDM pharmacists’ dose planning (n = 118, Figure S2 and Table S7). Input variables were selected based on the considered information while dose planning: the covariates for teicoplanin pharmacokinetics (age, BW, BMI, CL_CR_, and serum albumin level) and the timing of dose planning (day of the week [T1–T7] and time of the day [T0]). The latter comes from our practice of adjusting the dosing regimen, such that the initial TDM is performed on weekdays.

We divided eligible patients into a training group and a testing group at a 94:24 ratio (approximately 80:20). Subsequently, the neural network model to predict TDM pharmacists’ dose planning was trained on the patients in the training group. The prediction accuracies for the loading and maintenance doses (i.e., identical dosing as TDM pharmacists) were both 100% on the training dataset, indicating that the model learned how TDM pharmacists decide dosing regimens. However, in the testing trial, the model scored significantly lower prediction accuracies of 54.2% and 62.5% for the loading and maintenance doses, respectively (Figure 2A, B, left and Table S9). These results indicate that overfitting occurs, which is most likely attributable to the small datasets (17). Because the current dataset consisted of dose planning by multiple pharmacists, each with varying levels of expertise, possible heterogeneity in dose planning among pharmacists may also have contributed to the decrease in accuracy. Next, we retrospectively analyzed the target attainment by ML dose planning using the Bayesian method (2). The original target attainment rate in the testing group was 81.0% (17/21). Importantly, the rate was expected to slightly increase when ML was applied (95.3% [20/21], Figure 2C, left and Table 5). We hypothesized that ML complemented the weaknesses of suboptimal TDM pharmacists by learning policies from other pharmacists. Taken together, these results indicate that although dose planning tends to differ from that of TDM pharmacists, ML is competent for tailoring teicoplanin dosing as TDM pharmacists.

**Figure 2.**
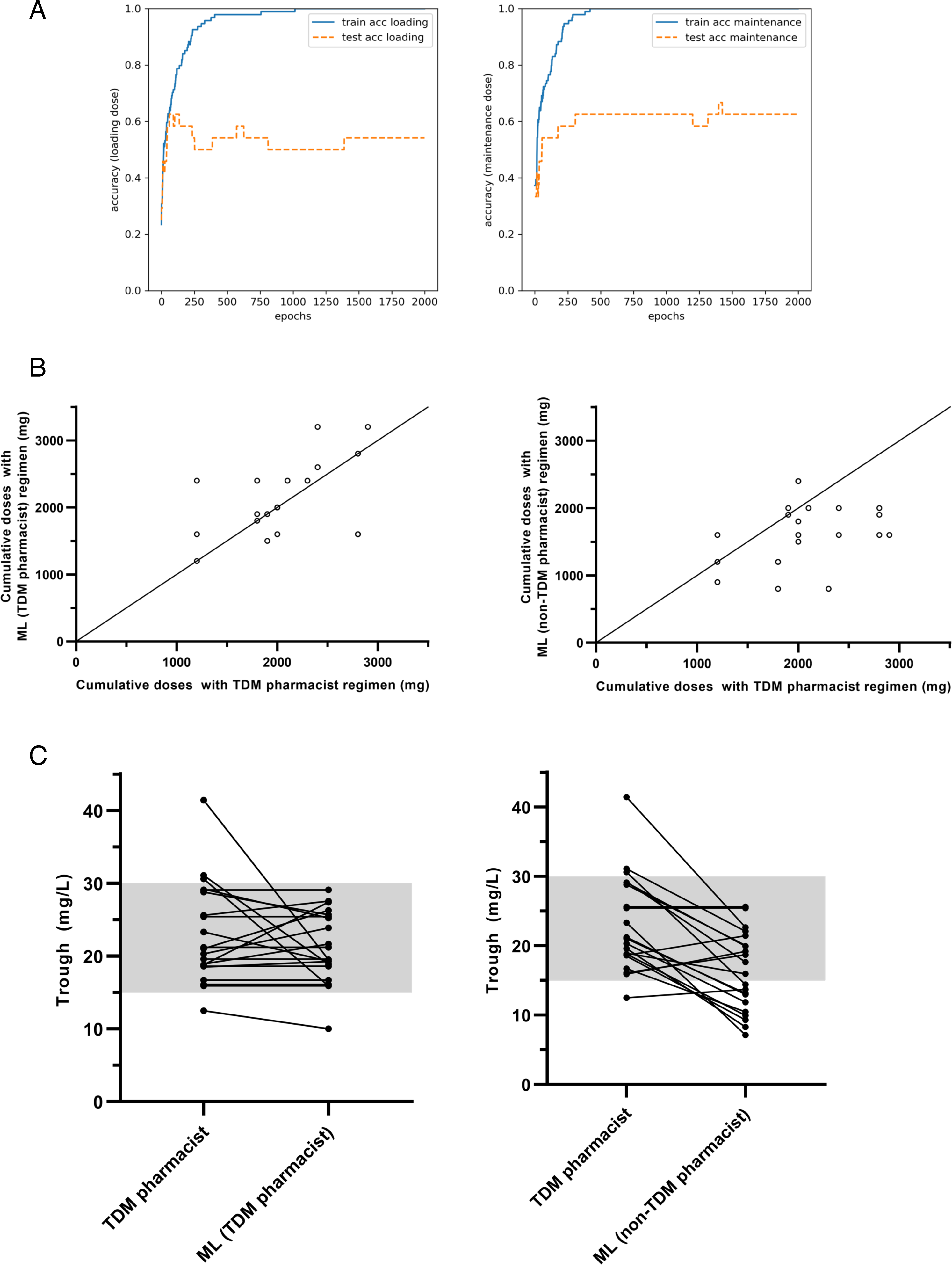
**(A)** Learning curves of the neural network through training and testing using dataset from the intervention group. **(B)** Cumulative dose of teicoplanin before initial TDM in TDM pharmacists and ML regimens. (Left) TDM pharmacists vs. ML trained with cases with intervention (TDM pharmacists ML). (Right) TDM pharmacists vs. ML trained with cases without intervention (non-TDM pharmacists ML). Line indicates reference line of y = x. **(C)** Serum concentration at initial blood sampling with TDM pharmacists and ML regimens. (Left) TDM pharmacists vs. ML trained with cases with intervention (TDM pharmacists ML). (Right) TDM pharmacists vs. ML trained with cases without intervention (non-TDM pharmacists ML). Gray area indicates therapeutic windows (15–30 mg/L).

**Table 5.** Target attainment (internal validation)

| Group<br>Trough (mg/L) | TDM pharmacists (%) | ML by TDM pharmacists (%) | ML by non-TDM<br>pharmacists (%) |
| --- | --- | --- | --- |
| <15 | 1 (4.8) | 1 (4.8) | 10 (47.6) |
| 15–30 | 17 (81.0) | 20 (95.3) | 11 (52.4) |
| >30 | 3 (14.3) | 0 (0.0) | 0 (0.0) |

Abbreviations: ML, machine learning.

As a control, we also trained the ML using dose planning by non-TDM pharmacists (i.e., the cases without intervention, Table S8) and then analyzed the target attainment by this ML. As expected from the poor target achievement (Table 2), ML trained on cases without intervention failed to maintain the target attainment of TDM pharmacists. The target attainment was estimated to decrease from 81.0% (17/21) to 52.4% (11/21) (Figure 2C, right, Table 5, and Tables S9 and S10). This decrease was attributed to the subtherapeutic doses of teicoplanin; the incidence of subtherapeutic exposure was estimated to increase from 4.8% (1/21) to 47.6% (10/21) (Figure 2B, right, Table 5, and Tables S9 and S10). This result mirrors the propensity of non-TDM pharmacists to underdose (Tables 2 and 4). Taken together, these results suggest that ML model training with the TDM pharmacists’ dataset augments model performance in tailoring the teicoplanin dose.

### 2.4 Evaluation of the ML on the external validation dataset

Finally, we investigated whether ML improved target trough attainment. Toward this end, we retrained the ML with whole dataset of TDM pharmacists’ dose planning (n = 118, Table S7) to improve the prediction. Subsequently, we applied retrained ML to patients in the nonintervention group without an ICU/HCU stay who underwent TDM (n = 81, Figure 1 and Table S8). For both loading and maintenance doses, the retrained ML scored 100% accuracy on the training dataset (Figure S3). We also calculated feature importance, which is proportional to the contribution of the features to the ML decision. For the loading dose, BW was the most important feature for the ML output (i.e., dose determination). For the maintenance dose, BW, BMI, and CL_CR_ represented major contributions to the ML output. For both loading and maintenance doses, the timing of dose planning (T0–T7) also contributed to the ML decision, which is reminiscent of our practice of adjusting dose planning to perform TDM on weekdays (Figure 3A). The small roles of T6 and T7 (dose planning on Saturday and Sunday, respectively) in dose planning were likely due to the lack of data (both n = 1) to optimize the weight parameters (Table S7).

**Figure 3.**
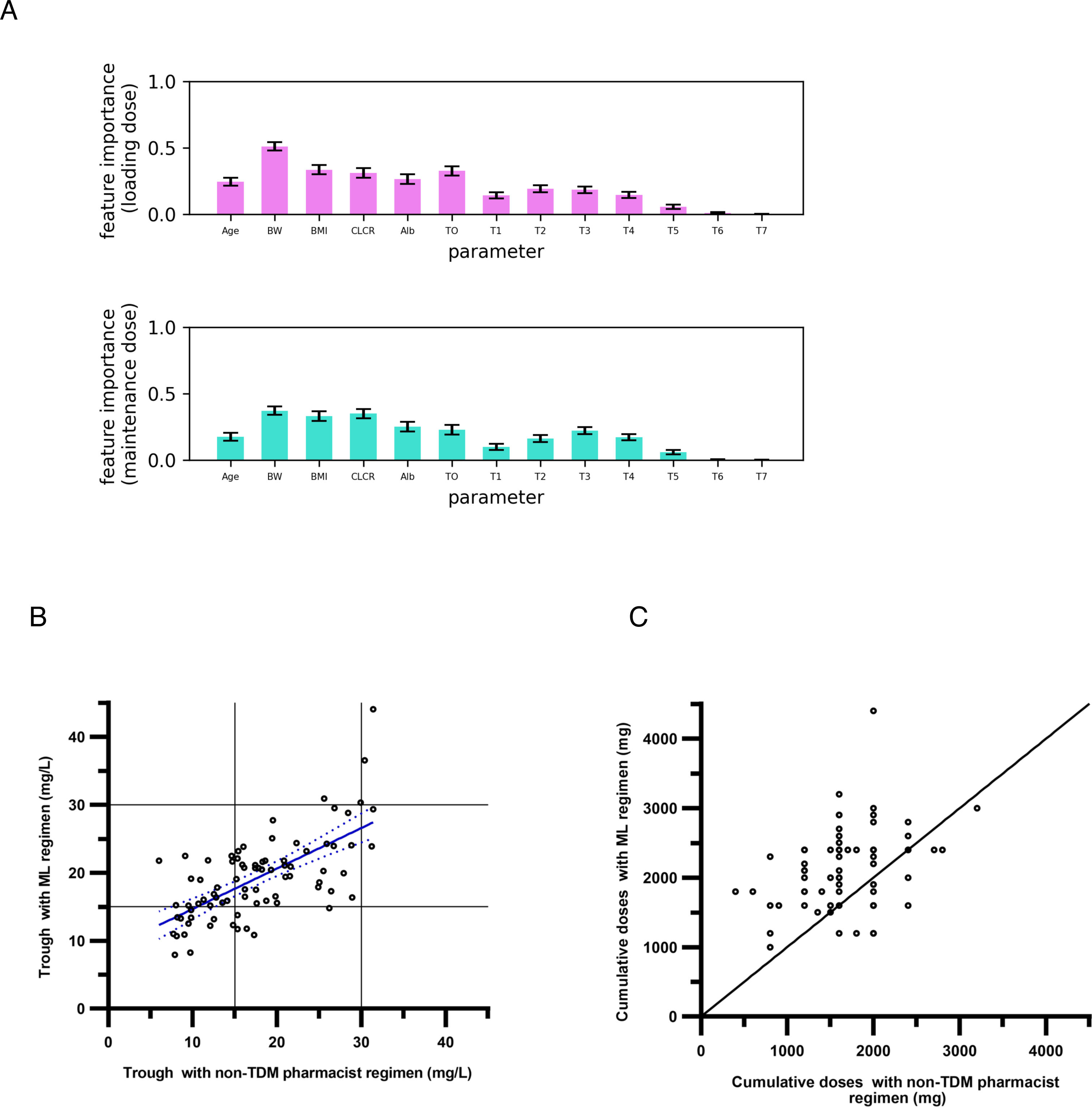
**(A)** Permutation feature importance for loading dose (top) and maintenance dose (bottom) decision. **(B)** Serum concentration at initial blood sampling with non-TDM pharmacists and ML regimens. Blue line and blue dashed lines indicate regression line and 95% confidence interval, respectively. **(C)** Cumulative dose of teicoplanin before initial TDM with non-TDM pharmacists and ML regimens. Line indicates reference line of y = x.

The target attainment rate in the nonintervention group was 58.0% (47/81) (Table 2). Notably, if ML dosing regimens were applied, target attainment rates increased from 58.0% to 72.8% (59/81, p = 0.047, Figure 3B and Table 6). This increase was mainly attributed to the enhanced dose regimen, which prevented subtherapeutic drug exposure; when applying the ML, subtherapeutic levels at initial TDM were expected to decrease from 37.0% (30/81) to 22.2% (18/81) (Figure 3C and Table 6). Meanwhile, notably, enhanced dose planning by ML was accompanied by an incidence of overexposure (e.g., case no. 182, Tables S11 and S12). Therefore, ML may increase the risk of adverse effects of teicoplanin, such as thrombocytopenia, which is observed when trough concentrations exceed 40 mg/L (2, 18, 19).

**Table 6.** Target attainment (external validation)

| Group<br>Trough (mg/L) | Nonintervention (%) | ML by TDM pharmacists (%) | p-value<br>(Chi-square test) |
| --- | --- | --- | --- |
| <15 | 30 (37.0) | 18 (22.2) | 0.047 |
| 15–30 | 47 (58.0) | 59 (72.8) |  |
| >30 | 4 (4.9) | 4 (4.9) |  |

Abbreviations: ML, machine learning.

Altogether, similar to the dose planning by the TDM pharmacists, the current ML is expected to play a role in tailoring dose planning, which contributes to early therapeutic exposure and consequently leads to treatment success. The model shown in Figure 3 is freely available at https://github.com/Matsuzaki-T/TEIC_study.git.

## 3. Discussion

A growing body of evidence has suggested that pharmacists’ intervention in initial dose planning leads to early adequate drug exposure, indicating the importance of pharmacists’ knowledge for tailoring initial dose planning (20, 21). Given the importance of pharmacists’ knowledge, we harnessed the power of ML techniques to integrate such knowledge into a predictive model. To validate this hitherto unexplored approach to expedite clinical decisions during the dose planning phase, we previously trained the ML model for vancomycin dose planning using a dataset of dose planning by pharmacists experienced in TDM. The results showed that the target attainment with the dosing regimen by the resultant ML was estimated to be the same as the pharmacists’ regimen, indicating that the ML was as competent in tailoring dose planning as well-trained pharmacists (16). This result motivated us to develop a predictive model for another clinically important antibiotic, teicoplanin, which is equally effective but better tolerated than vancomycin, with a lower risk of nephrotoxicity (22, 23).

In this study, we first validated the dose planning for teicoplanin by TDM pharmacists in our hospital (Tables 1, 2, and 4). Compared with vancomycin, evidence that pharmacists’ intervention improves clinical outcomes of teicoplanin treatment is limited. One study demonstrated that pharmacists’ intervention improved the attainment of targeted teicoplanin concentrations (24). However, in this study, target achievement was recorded whenever concentrations reached the therapeutic window during treatment. Therefore, whether pharmacists intervention improved early target attainment was unclear. Although the increase in target attainment rate was limited to patients without an ICU/HCU stay, the current study marks the first time that dose planning of teicoplanin by experienced pharmacists led to the achievement of therapeutic windows accompanied by increased adherence to appropriate blood sampling (Tables 1 and 2). These results, along with similar outcomes reported for vancomycin (20, 21), indicate the clinical significance of pharmacists’ (especially pharmacists experienced in TDM) intervention in the treatment of MRSA infection.

The feature importance of the current ML was in accordance with the conventionally employed predictive covariates for dosing (Figure 3A) (9, 25). The timing of dose planning also played a role in the current ML, indicating that our practice of adjusting dose planning in a time-dependent manner was incorporated into the ML.

The prediction accuracy of ML was suboptimal at 54.2% and 62.5% for the loading and maintenance doses, respectively (Figure 2A). In our previous study, ML for vancomycin initial dose planning also showed limited prediction accuracies of 59.1% and 68.2% for the loading and maintenance doses, respectively (16), indicating overfitting. As small datasets are susceptible to overfitting, ML training with larger sample sizes is required (17). The simple architecture of the current neural network, potential heterogeneity in dose planning among TDM pharmacists, and intrinsic difficulty of multiclass classification tasks may have also affected the prediction accuracy (17). Nevertheless, in line with our previous study, ML predicted the dose planning that was expected to reach the target trough levels as the original regimens by pharmacists (Figure 2C and Table 5) (16). These results suggest that the ML learns basic policies for tailoring the teicoplanin dose in a supervised manner. Interestingly, in contrast to the ML trained with dose planning by TDM pharmacists, the ML trained with dose planning without intervention failed to maintain target attainment by TDM pharmacists (Figure 2C and Table 5). The number of cases with subtherapeutic exposure was expected to increase from 4.8% (1/21) to 47.6% (10/21) when ML was applied, mirroring the propensity toward underdosing in cases without intervention (Tables 2 and 4). Taken together, these results suggest that imitation learning can be used to tailor antibiotic doses.

We also tested an external validation cohort using cases without intervention. The results showed that the target attainment rates increased when applying ML trained with dose planning by TDM pharmacists (Figure 3B and Table 6). This increase was mainly a result of the enhanced loading doses of ML, as cases with subtherapeutic exposure decreased from 37.0% (30/81) to 22.2% (18/81) (Figure 3B and Table 6). These results indicate that ML plays a role in individualizing the initial dosing regimen as TDM pharmacists. Meanwhile, enhanced loading doses by ML were associated with the incidence of overexposure (e.g., case no. 182, Tables S11 and S12). Therefore, at this preliminary stage, the current model should be used with caution. Moreover, notably, the trough concentrations of the ML regimen were estimated using Bayesian prediction accompanied by assumptions regarding the timing of drug administration and blood sampling, which harbors several limitations in evaluating the performance of the current model. The generalizability and clinical utility of this model should be rigorously evaluated in future prospective studies.

In addition to the abovementioned limitations, there are a few other limitations to be noted. First, the current predictive model was derived from a single center with a small sample size, thereby lacking external generalizability. Second, the dose planning by TDM pharmacists in the current study targeted trough levels of 15–30 mg/L; however, recent guidelines have suggested trough levels of 20–40 mg/L for serious and/or complicated MRSA infections (2). Third, although pathophysiological conditions (e.g., hematological malignancy) have been reported to affect the pharmacokinetics of teicoplanin, we did not employ these factors in the ML training phase (26, 27). The next step for developing a more sophisticated ML model for tailoring teicoplanin dosing is to use this information in ML training. Finally, the majority of the study participants were older adults, which may have influenced the prediction performance in younger adults (Table 3).

In conclusion, we validated an ML approach to develop a model for tailoring the initial dosing regimen of teicoplanin, which holds promise for complementary dose planning by clinicians. This study, together with our previous work on vancomycin (16), provides a new avenue for achieving early target exposure, which has important ramifications for the successful treatment of invasive MRSA infections.

## 4. Materials and methods

### 4.1 Study participants

This was a single-center, retrospective, observational study of hospitalized patients who received intravenous teicoplanin between August 2019 and April 2022 at Nagoya University Hospital. TDM pharmacists were defined as pharmacists dedicated to TDM. Patients who commenced teicoplanin treatment with TDM expert-recommended dose regimens during the study period were also included. The exclusion criteria were as follows: patients aged <18 years; undergoing peritoneal dialysis or hemodialysis (including continuous hemodiafiltration); undergoing ECMO; receiving intervention by experts after the initial dose; restarting teicoplanin treatment within 7 days; who were immobile, which indicates decreased muscle mass and may cause overestimation of CL_CR_; with surgical antibiotic prophylaxis; and with missing data on sex, age, BW, BMI, and serum albumin and creatinine levels.

### 4.2 Comparison of appropriate blood samplings and trough concentrations between the intervention and nonintervention groups

Eligible patients with serum teicoplanin concentrations were enrolled to evaluate the dose planning by TDM pharmacists. In this analysis, the patients were divided according to whether they received dose planning by TDM pharmacists.

Patients with intervention were excluded if the time of blood sampling was not described, protocol deviation occurred, dosing regimen was changed by TDM pharmacists after the initial dose, or blood samples were collected after the completion of teicoplanin treatment. Because blood samples for monitoring teicoplanin trough concentrations should be collected at least 18 h after the last dose (2), we monitored the timing of blood sampling. In cases where initial TDM was performed at the recommended time (18 h after the last dose), the target trough achievement (15–30 mg/L) in the intervention group was compared with that in the nonintervention group.

Propensity scores were calculated using logistic modeling, with TDM pharmacists’ dose planning as the dependent variable. Independent variables included age, BW, BMI, serum albumin level, CL_CR_ calculated using the Cockcroft–Gault equation (28), and timing of dose planning, which were used as input variables in ML construction. The patients were matched 1:1 using the nearest-neighbor technique, with a caliper distance limited to 10% of the standard deviation of the pooled propensity scores.

### 4.3 Building of the neural network model

The dataset used in this study included clinical and routine laboratory data, initial dosing regimens, and serum teicoplanin concentration at initial TDM (if measured). Age, BW, BMI, serum albumin level, and CL_CR_ were used as features to predict the initial dosing regimens (loading and maintenance doses). We also used the timing of the dose planning (day of the week [T1–T7] and times of day [T0]) as input variables because TDM pharmacists consider dosing regimens so that initial TDM is performed on weekdays as possible. The dataset (n = 118) was divided into training and test datasets in a 94:24 ratio (approximately 80:20). Numeric input variables (age, BW, BMI, serum albumin level, CL_CR_, and time of day [T0]) in the training data were normalized using the following equation:

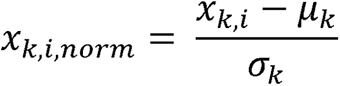

where *x_k,i_* is a value of parameter *k* in sample *i*, *x_k,i,norm_* is a normalized value of parameter *k* in sample *i*, *µ_k_* is a mean of parameter *k*, and *σ_k_* is a standard deviation of parameter *k*.

We applied the same scaling to the input variables in the testing data as in the training data to avoid shifting the distribution of the data. For ICU/HCU stay, the variable takes 1 for ICU/HCU stay and 0 otherwise. On each day of the week, the variables were binarized using one-hot encoding, where T1, T2, T3, T4, T5, T6, and T7 represented Monday, Tuesday, Wednesday, Thursday, Friday, Saturday, and Sunday, respectively. Output variables (loading and maintenance doses) were binarized using one-hot encoding.

We built a two-layer neural network model for dose planning, as described elsewhere, using Python (29, 30). The structures of the network for the loading and maintenance doses were the same. Briefly, the network was composed of an input layer, a hidden layer, and an output layer (Figure S2). The activation functions in the hidden and output layers were both sigmoid. The number of hidden neurons was set to 15, which was determined by the sum of 2/3 of the size of the input layer neurons (n = 13) and output layer neurons for the maintenance dose (n = 7), as previously proposed (31). Because adding the number of output layer neurons to the loading dose (n = 14) did not result in increased predictive accuracy (data not shown), we set the same number of hidden neurons in both ML.

We trained the neural network to minimize empirical loss over the training data. In this study, we employed cross-entropy loss as the loss function *L*, which was parameterized by the weight matrices *W1*, *W2*, *b1*, and *b2* (Figure S2):

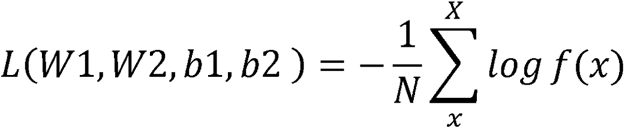

where *X* is a matrix of the input variables from each training dataset *x*, *N* indicates the number of training data points, and *f*(*x*) indicates the score for the correct class.

We optimized each parameter in the model using stochastic gradient descent with an adaptive learning rate using AdaGrad (32):

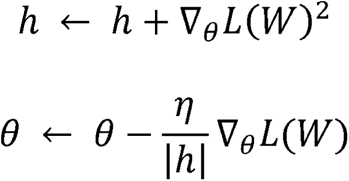

where *θ* is a parameter of weight matrix and *η* is a learning rate (in this study, 0.1).

### 4.4 Feature importance analysis

In the training process, the permutation feature importance (Breiman–Cutler importance) was calculated, as described in previous studies (33, 34). In this process, a single feature value was randomly shuffled while keeping the other input variables constant. Subsequently, decreases in the prediction accuracy, which indicates how the feature contributes to the M decision, were collected for each input variable. We repeated this process 20 times to measure the mean decrease in the prediction accuracy for each feature:

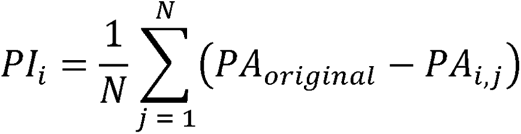

where *PI_i_*(permutation importance) indicates the permutation importance of feature *i*, N is the number of repetitions (in this study, 20), *PA_original_* indicates the original prediction accuracy, and *PA_i,j_* indicates the prediction accuracy upon shuffling feature *i*.

### 4.5 Estimation of trough concentration with the regimen using the ML model

Data, including serum teicoplanin concentrations at appropriate time points (18 h after the last dose), were included in this analysis. If the ML dosing regimen by the ML was identical to the original dosing regimen, the measured serum teicoplanin concentration was defined as the trough concentration with the ML dosing regimen. Otherwise, the serum teicoplanin concentration in the ML regimen was estimated using Bayesian estimation under the following assumptions:

(i) If the cumulative number of doses was the same in the original and ML regimens, only the dose was changed.
(ii) If the ML recommended an additional loading dose, additional loading doses were added 8 h after the last dose and every 8 h thereafter.
(iii) If the ML did not recommend a maintenance dose, blood sampling was performed 18 h after the last dose.
(iv) If the ML recommended maintenance dose, every 24 h maintenance dose was assumed to be administered at 9:00 a.m. (at least 8 h after the last dose), followed by TDM sampling at 9:00 a.m. the next day.

### 4.6 Statistical analyses

We used the Mann–Whitney U test to evaluate continuous data. For categorical data, we used Fisher’s exact test or the chi-squared test. All statistical tests were two-tailed, and p values < 0.05 were considered statistically significant. Statistical analyses were performed using the Python Statistics module.

## Data Availability

All data produced in the present study are available upon reasonable request to the authors.

https://github.com/Matsuzaki-T/TEIC-AI

## 5. Supplementary material

Supplemental material is available online only.

The code for the predictive model with pretrained weights and modules reproducing the results of this study is available at https://github.com/Matsuzaki-T/TEIC_study.git.

## 6. Ethics approval

This study was conducted with the approval from the Ethic Committee of Nagoya University Hospital (Approval No. 2022-0071).

## 7. Acknowledgements

This work was supported by Morinomiyako Medical Research Foundation, the JSPS KAKENHI (Grant Numbers JP 20H03428 and 22K17824), and the Research Funding for Longevity Sciences (22–21) from National Center for Geriatrics and Gerontology (NCGG), Japan.

