## Supplemental Figure 1 to 3 for "Integration of pharmacists’ knowledge into a predictive model for teicoplanin dose planning"

Figure S1

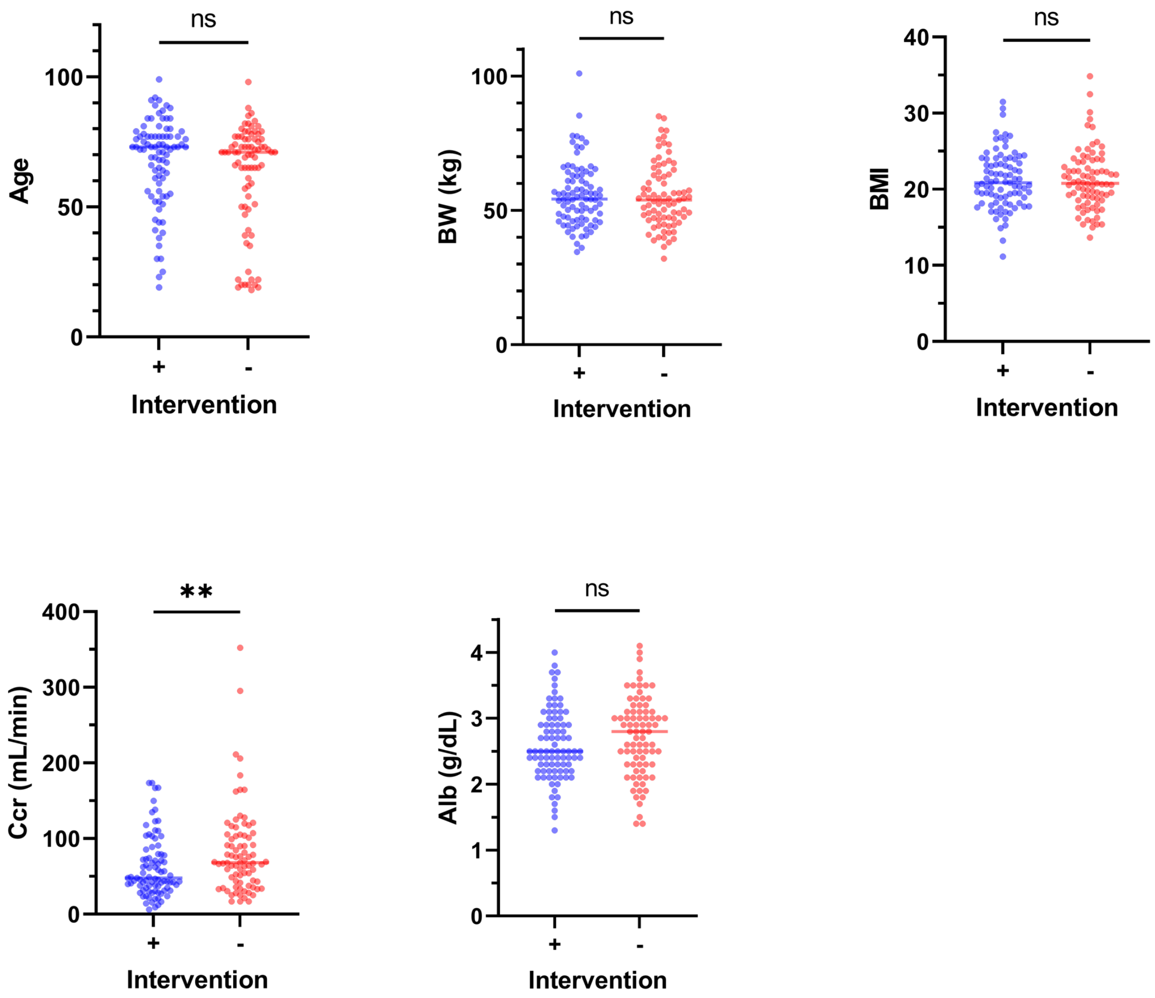


**Figure S1.** Baseline characteristics of patients without ICU/HCU stay included in the analysis of target attainment (**p <0.01, ns. not significant). See also Table 3.

Figure S2

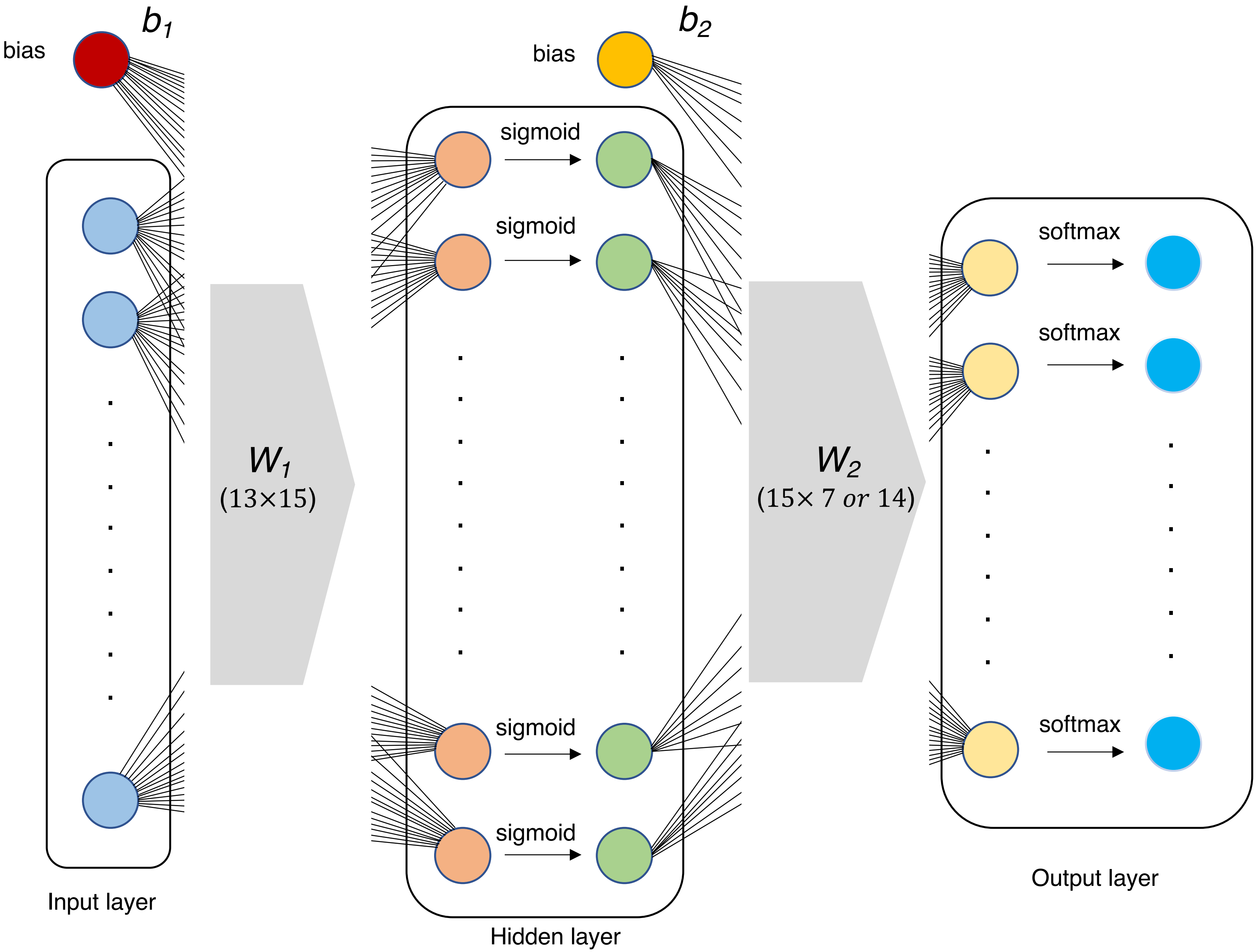


**Figure S2**. The architecture of the predictive model in this study.

Figure S3

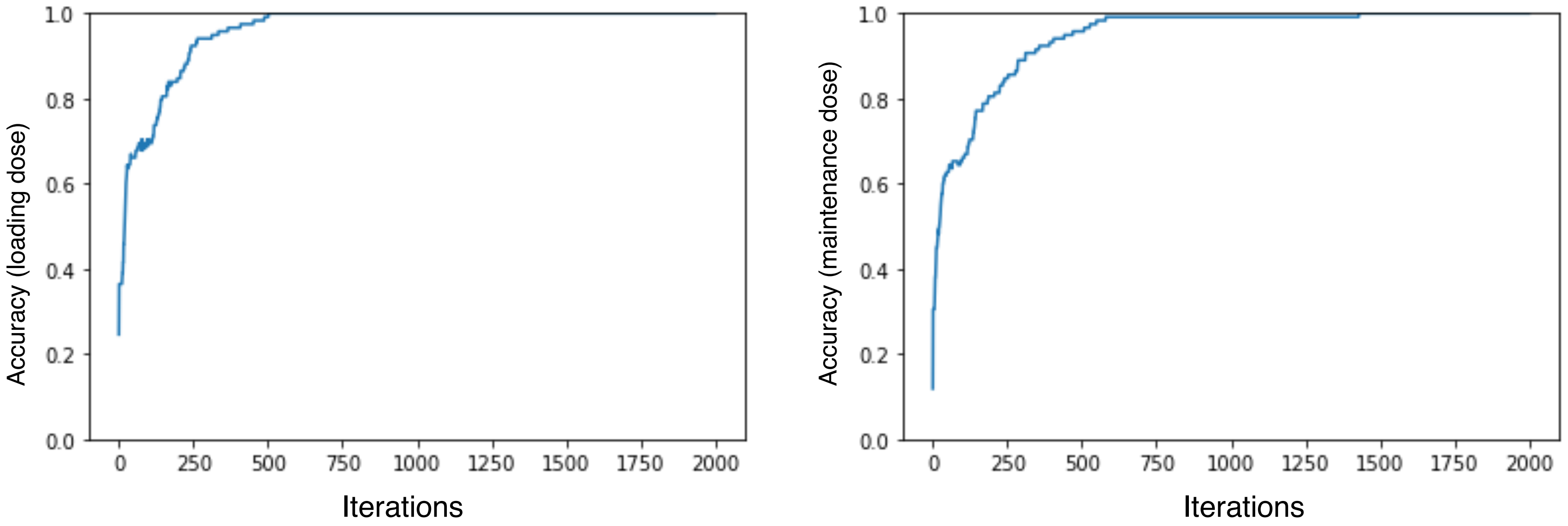


**Figure S3**. Learning curves of the neural network in Figure 3.
